# Trigeminal Nerve Stimulation (TNS) for Children with Attention Deficit/Hyperactivity Disorder and Fetal Alcohol Spectrum Disorder: Feasibility Study Protocol

**DOI:** 10.1101/2025.07.07.25331025

**Authors:** Joseph O’Neill, Shantanu Joshi, Jeffry R Alger, Benjamin N Schneider, Mary J O’Connor

## Abstract

Symptoms of attention deficit/hyperactivity disorder (ADHD) are common, severe and highly impairing in children with prenatal alcohol exposure (PAE), but often non-responsive to medication, leaving many with no beneficial treatment. External trigeminal nerve stimulation (TNS) is a minimal risk, non-invasive neuromodulatory intervention that is FDA-cleared for ADHD. No formal trial, however, has tested TNS in children with known PAE. We present here the protocol of the first clinical trial of TNS in children with ADHD associated with PAE. The study also uses multimodal MRI to explore possible brain mechanisms of TNS. An open-label pilot will be conducted of 4 weeks of TNS in 30 youth with ADHD associated with PAE, recruited in Southern California. Children will receive TNS nightly for 4 weeks. Safety, tolerability, and preliminary efficacy will be evaluated. Efficacy outcomes include change in the physician-administered parent-rated ADHD Rating Scale and the Clinical Global Impression Improvement (CGI-I) score for ADHD. Effects on executive function, mood, and sleep are also assessed. Maintenance of effects is evaluated at 4-week follow-up. TNS-related brain changes and predictors of response are measured using structural MRI, diffusion tensor imaging (DTI), magnetic resonance spectroscopy (MRS), and resting-state fMRI before and after treatment. This study will determine whether TNS is feasible in children with ADHD and PAE, whether it improves clinical and cognition symptoms, and whether efficacy persists 4 weeks. Trial registration: clinicaltrials.gov (NCT06847165) and protocol ID: IRB-24-0648-AM-007 June 16, 2025.

## Introduction

Despite decades of efforts to curb ethanol consumption in pregnancy, prenatal alcohol exposure (PAE) still affects up to 5% of US children [1]. High disability arises from ADHD-like behavioral symptoms, including hyperactivity, impulsivity, and executive function deficits [2-4], that occur in 50-95% of children with PAE [5-7]. In ADHD without PAE, these symptoms often improve spontaneously by adulthood [8-9]. But in individuals with PAE they typically persist and evolve into problems in behavioral and emotional regulation and adaptive function [7,10-11] leading to “secondary disabilities” [10,12], including school failure, legal trouble, substance abuse, mental illness, and suicide [7,12-15]. Secondary disabilities add to the >$4 billion/year ([16]; NIAAA estimate) public health cost of PAE. Unfortunately, controlled treatment studies designed to address these disabilities are rare [17]. Most commonly, psychostimulants are prescribed for behavioral symptoms in ADHD both with and without PAE [18], but while these drugs lead to routine improvement in idiopathic ADHD, their efficacy in ADHD with PAE is *not* well established [19]. Rather, there is much evidence that stimulants are less effective for ADHD with PAE [20-23]. A small retrospective study recommending amphetamines for PAE [24] remains unreplicated and non-response of individuals with ADHD to stimulants remains a possible red flag for PAE [11]. Side effects to stimulants, moreover, may occur at higher rates in PAE [25-27]. While promising evidence-based behavioral interventions for PAE have emerged at our Center and elsewhere [28-34], children with PAE often fail to retain gains achieved with behavioral therapies.

Trigeminal nerve stimulation (TNS; [35]) was recently found safe and efficacious for core (inattention, hyperactivity) and executive-function symptoms of ADHD in children in an open pilot and a sham-controlled double-blind randomized controlled trial (RCT) at UCLA [36-37]. The response rate (50%) was appreciable for psychiatry, comparable to the 40-60% efficacy of SRIs for major depressive disorder (MDD; [38]). While these trials led to FDA-clearance of TNS for pediatric ADHD, patients in those trials were *not* screened for PAE. Given the symptom overlap of PAE and ADHD and the fact that certain putative sites of action of TNS in the brain match sites where we found imaging abnormalities in PAE [39-41], we are optimistic that TNS will also be safe and effective in alleviating the behavioral symptoms of children with ADHD and PAE. But formal testing of this expectation is required if we are to have a firm basis for recommending TNS as a therapy. Our data acquired with magnetic resonance (MR) neuroimaging have documented differences between children with ADHD symptoms with and without PAE in many brain regions [42]. Such differences may influence clinical response to TNS. A clinical trial with imaging may answer this question and may help elucidate the therapeutic mechanism of TNS, which is mostly unknown for the many conditions it treats.

If TNS is safe and efficacious for children with ADHD with PAE, this research will yield a much needed new, FDA-cleared nonmedication therapy for managing PAE. Our imaging studies could, moreover, improve TNS by uncovering where it acts in the brain, which imaging measures are changed by it, and which portend good response. If, in contrast, TNS is not safe or effective for PAE, it will still be impactful to demonstrate to clinicians that PAE is a negative prognostic factor when treating patients with TNS. Since PAE is highly underdiagnosed or misdiagnosed [1,7,43-44], screening for PAE could then raise the efficacy of TNS for ADHD.

This research is associated with NIAAA grant R61AA031029/R33 (Principal Investigator O’Neill). This pilot grant will determine whether TNS is feasible for PAE: Do children with PAE comply with TNS procedures? Does TNS have any serious side effects in these children? Is there an indication that TNS relieves symptoms of PAE? Is such relief retained in follow-up? If the pilot shows feasibility, we will apply for continued funding of an R33 phase, a double-blind sham-controlled randomized clinical trial to determine whether TNS is truly efficacious for children with PAE. This paper aims to address an urgent unmet need by first describing the protocol for the pilot phase of the study designed to test the feasibility, tolerability, compliance, and adverse effects associated with TNS treatment. We will also assess whether children show diminished hyperactivity/impulsivity, increased attention, improved executive function, and emotional control.

## Materials and Methods

### Study design and setting

The SPIRIT Schedule of enrollment, interventions, and assessments appears in Fig 1. This is an open-label feasibility study of 28 days of nightly TNS treatment in 30 pediatric outpatients with ADHD and PAE. Each 8-hr treatment session is delivered overnight in sleep. Participants receive neuropsychological assessments and multimodal MR brain imaging before and after the treatment regimen. The trial is being conducted at the Division of Child & Adolescent Psychiatry Clinical Trials Hub, Semel Institute for Neuroscience, David Geffen School of Medicine at UCLA, Los Angeles. All multimodal MR scans are completed on the 3-T Siemens Prisma MRI scanner at the UCLA Staglin Center for Cognitive Neuroscience (CCN) at the Institute. Recruitment began on April 1, 2025, and is expected to continue until September 2026.

**Fig 1.**
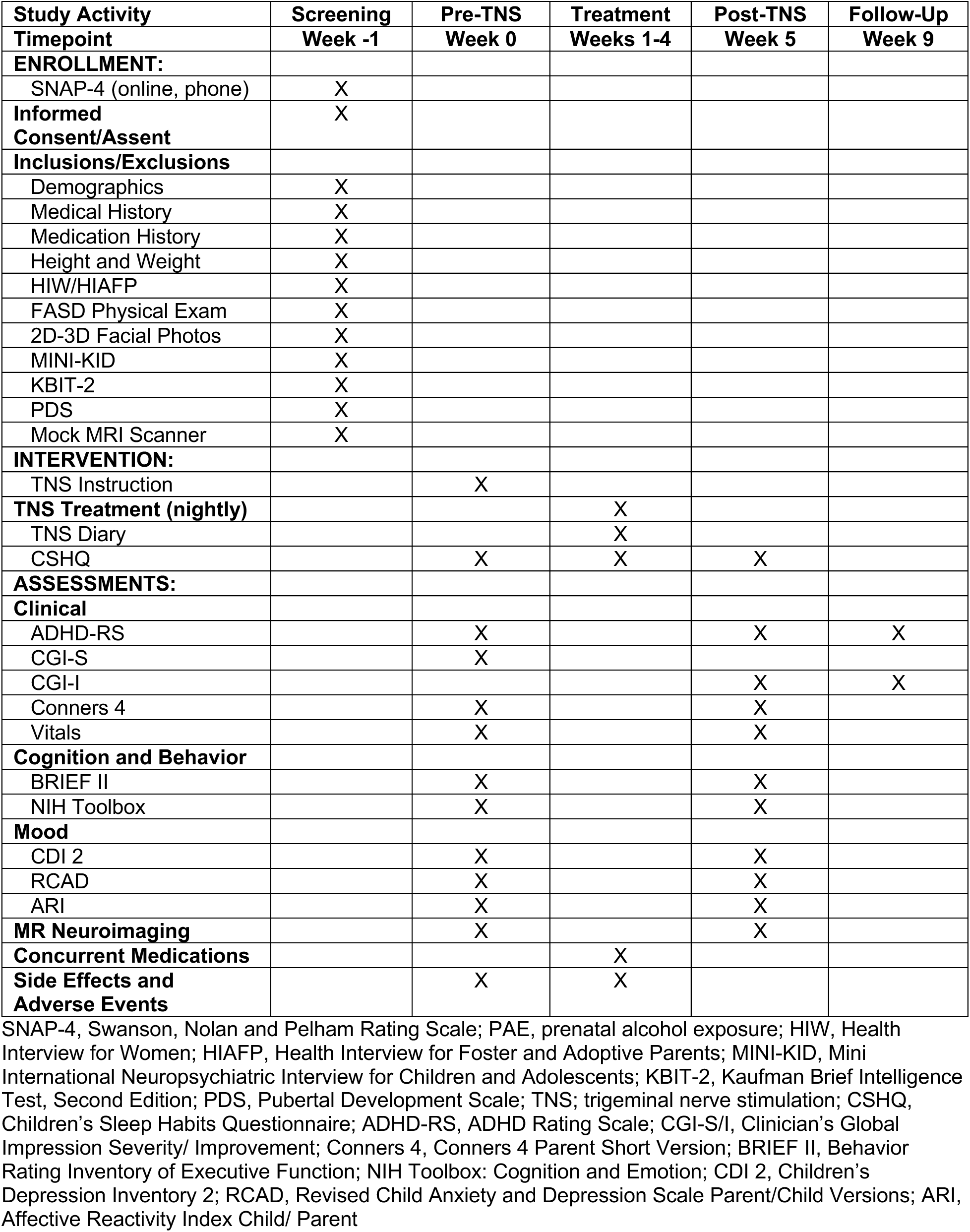
SPIRIT Schedule of enrollment, interventions, and assessments.

### Participants

Children admitted to the study will be between 8 and 12 years of age with a history of ADHD with PAE. Primary caregivers will be referred to as “parents” with the understanding that they could be biological, foster, or adoptive parents. The study team will use a checklist of Inclusion/Exclusion criteria when making the enrollment decision for each participant. The team will also maintain a CONSORT-type diagram (Supplemental Materials) of participant screenings, exclusion, enrollment, retention, and dropout throughout the study. Data will specify reasons for study exclusion and dropout.

#### Participant identification and recruitment; Sample size

Participants are recruited through the Child and Adolescent General Outpatient Clinic, the Child Day-Treatment Program, and the Psychopharmacology Clinic in the Semel Institute. Participants are recruited from the UCLA Division of General Pediatrics at Mattel Children’s Hospital. Beyond UCLA, we recruit from practitioner referrals, schools, and the community using flyers posted at YMCAs, afterschool programs, and parks and recreation programs, as well as from FASD United, and FASD of Southern California parent support groups. Social media notify families of the study, for this we use Meta Ads. We expect to enroll 30 patients (24 post-attrition) over 2 years. This sample size was calculated from a power analysis using effect sizes from [37] and an attrition rate of 20% based on our prior studies of PAE.

##### Inclusion criteria

(1) Children aged 8-12 years at study entry; (2) Fetal alcohol syndrome, partial fetal alcohol syndrome, or alcohol-related neurodevelopmental disorder per modified Institute of Medicine criteria [45]; (3) PAE >6 drinks/week for ≥2 weeks and/or 23 drinks on 22 occasions throughout gestation per Health Interview for Women/Health Interview for Adoptive and Foster Parents (HIW/HIAFP; [46]); (4) Scores >13 on both Inattention (questions 1-9) and Hyperactivity (questions 10-18) of the SNAP IV; (5) ADHD Diagnosis (DSM-5) based on the MINI-KID Parent Version 7.0.2 “ADHD Current-Yes” on the ADHD Module; (6) Full-Scale IQ >70 on the Kaufman Brief Scale of Intelligence 2 (KBIT 2); (7) T scores >65 on Inattention, Hyperactivity/Impulsivity, and Total Score on the Conners 4 Parent Short Form; (8) Parent and child able to complete testing in English; (9) Child able to cooperate during MRI; (10) Child able to comply with study procedures.

##### Exclusion criteria

(1) Active suicidal ideation as evidenced by meeting criteria for “Current” Suicidality on the Suicide Behavior Disorder module of the MINI KID Parent Version 7.0.2; (2) Comorbidity with other major psychiatric disorders except conduct disorder, oppositional defiant disorder, anxiety, and depression (often associated with ADHD with PAE) as measured by the MINI KID Parent Version 7.0.2; (3) Other toxic exposures per HIW/HIAFP whose influence clearly surpasses that of alcohol (very rare) per study physician judgement; (4) Known genetic syndrome associated with ADHD-like symptoms including fragile X, tuberous sclerosis, or generalized resistance to thyroid hormone; (5) Serious medical or neurologic illness likely to influence brain function, e.g., seizures, closed-head trauma; (6) Gestation <34 weeks; (7) Ferromagnetic metal, claustrophobia, or other MRI or eTNS contraindication (e.g., insulin pumps or other body-worn devices).

### Eligibility Measures

#### Pre-Screening: SNAP-IV

The Swanson, Nolan and Pelham Rating Scale (SNAP IV; [47]) is a widely used scale with good validity and reliability for detecting ADHD and quantifying ADHD symptom severity in youth. We administer the SNAP-IV during online or telephone screening to a parent of each prospective enrollee. The purpose is to detect potential ADHD in the child and to screen out children with negligible ADHD symptoms. We also pre-screen for other inclusion and exclusion criteria, including possible PAE. Children likely to qualify are invited in for an in-person Eligibility Assessment at UCLA. Data from participants excluded at screening are fully anonymized; personal health information from these participants is destroyed.

#### Ethical considerations: Informed Consent/Assent

All procedures are performed in accordance with the 1964 Declaration of Helsinki and its amendments. The study was approved by the UCLA Medical Institutional Review Board (MIRB). Parents and children give written informed consent/assent prior to study inclusion. Consent/assent are obtained by authorized members of the investigative team. The study has a three-member Data Safety and Monitoring Board (DSMB) containing experts on PAE, clinical trials methodology, and statistics. The DSMB is independent of the study sponsor and has no competing interests. Its charter is available from the PI. Additionally, the study is subject to a mid-trial audit that is independent of the investigators and the sponsor. Important protocol modifications are reported to trial participants.

### Eligibility Measures

#### Demographics, Medical History, Current Medications

The Demographic Questionnaire measures common population characteristics including age, sex, ethnicity, parent education, family income, etc. The Medical History and Medication Questionnaires recall medical events experienced by the child as well as past and current medications, respectively.

#### Height and Weight

We assess whether these are affected by TNS treatment. In addition, height and weight percentiles allow measurement of the criterion for growth retardation in children with FASD. These measurements are entered as part of the PAE Physical Examination during the Eligibility visit as well as at the Pre- and Post-TNS Assessments.

#### Diagnosis of FASD: Prenatal Alcohol Exposure

Alcohol exposure is assessed using the Health Interview for Women (HIW) or the Health Interview for Adoptive and Foster Parents (HIAF)[46]. The HIW assesses frequency and quantity of typical and binge drinking and use of other teratogens prior to and following recognition of pregnancy. For adopted/fostered participants, information on prenatal exposure to alcohol and other teratogens is obtained *via* birth, medical, or adoption records or reports by reliable informants. Because many individuals with FASD are adopted or fostered, it is often necessary to use such records, and this is an accepted method in the scientific community for establishing PAE [48]. Criteria for alcohol exposure reported by the biological mother include >6 drinks/week for ≥2 week and/or ≥3 drinks on ≥2 occasions including the time periods prior to and following pregnancy recognition [45]. These criteria are based on findings that 1 drink/day (or >6 drinks/week) is an adequate measure of exposure for a FASD, and on epidemiologic studies demonstrating adverse fetal effects of episodic drinking of ≥3 drinks per occasion [45]. No child in the alcohol exposed group is accepted without a clear history of PAE.

#### Diagnosis of FASD: FASD Physical Exam

Every child is assessed for diagnostic features of FASD using the modified Institute of Medicine (IOM) criteria according to the updated Clinical Guidelines for Diagnosing Fetal Alcohol Spectrum Disorders [45]. This system examines the expression of four key diagnostic features of FAS: (1) growth retardation; (2) the FAS facial phenotype, including a flat upper vermillion border, flat philtrum, and short palpebral fissures; (3) neurodevelopmental dysfunction; and (4) gestational alcohol exposure. Growth retardation is defined as height and/or weight at or below the 10^th^ percentile on national growth charts at any point in time from birth. With regard to facial features, two of the three seminal facial features of FAS are required to be present. The upper vermillion border and philtrum are scored using a racially normed Lip-Philtrum Guide with scores of 4 or 5 meeting criteria consistent with an FASD [49]). Palpebral fissure length is scored using the Canadian palpebral fissure guidelines [50] and the guidelines for Black/African American and Hispanic children from Iosub et al. [51].

#### 3D Facial Photos

We take three 3D pictures of the face from the front, side, and at a ¾ angle. There are established links between neurodevelopment and the FASD facial phenotype. Without prior knowledge of PAE, FAS diagnosis [45,49] relies on accurate identification of abnormal facial features, growth deficits, and CNS problems. However, since the extent of facial abnormalities may be dose-dependent and more evident in severe cases or in older children, more effective quantification of facial dysmorphology, especially for mild or moderate cases and at younger ages may be needed. We use 3D facial photographs to objectively assess the potential in predicting the association among face and brain function and responses to TNS treatment. The 3D photos are supported by conventional 2D photos.

#### Diagnosis of ADHD and Co-Morbidities: MINI-KID

The Mini International Neuropsychiatric Interview for Children and Adolescents Parent Version (MINI-KID Parent Version 7.0.2; [52-53] is a structured diagnostic interview used to assess various mental health disorders in children and adolescents 6-17 years old. Questions are asked directly to the child’s parent, providing insight into their observations of the child’s behavior and potential symptoms. This allows for a more comprehensive evaluation compared to interviewing the child alone. Responses to the MINI-KID are binary (yes/no), indicating presence or absence of symptoms of a psychiatric disorder. We use the MINI-KID to establish diagnoses of ADHD and (exclusionary and non-exclusionary) psychiatric co-morbidities. The MINI-KID also has a section on suicidality. Positive responses including suicidal ideation with a plan make the child ineligible for the study and trigger our suicide prevention plan. The MINI-KID has previously demonstrated strong psychometric properties in clinical and general populations [52,54].

#### IQ Assessment: Kaufman Brief Intelligence Test, Second Edition Revised (K-BIT-2; [55])

The K-BIT-2 is a brief screening tool used to assess intellectual functioning in individuals 4-90 years old. The KBIT-2 yields 3 standard scores--Verbal, Nonverbal and IQ Composite Score. Each is an estimate of general intellectual functioning with mean of 100 and standard deviation of 15. The IQ Composite Score has high internal consistency across ages 4-18 (M = 0.93) with test-retest reliability of 0.88. The correlation between the IQ Composite score and the General Ability Index of the WISC-IV is 0.84. Purpose is to rule-out children with intellectual disability (IQ <70).

#### Pubertal Development Scale (PDS; [56])

Given the long-observed secular trend towards earlier puberty in children, we use the PDS to measure pubertal development as a covariate. It is a physically non-invasive test. Given that most youth are unreliable completing this questionnaire, it is advised to administer the scale to parents. The PDS includes three items that ask about growth in height, body hair, and skin changes. The PDS also asks about breast development and ministration in girls and deepening of voice and growth of hair on face in boys. Participants rate each item on a 1 (barely started) to 4 (seems complete) scale. The PDS demonstrates good internal consistency with Cronbach’s α ranging between 0.91 and 0.96 and high test-retest reliability (ICC=0.81–0.92).

#### Mock MRI Scanner Exposure

Children undergo standardized exposure at the mock scanner at the CCN. Ability to remain still during prolonged MRI studies and possible discomfort in the tight space of the scanner are assessed. Children unlikely to tolerate MRI scans without generating movement artifacts or with possible claustrophobia may be excluded from the MRI portion of the study.

### Trigeminal Nerve Stimulation (TNS) Treatment

#### TNS Instruction

Instructions on the use of the TNS study device are provided to each family along with a chance to practice using the instrument before treatment begins.

#### Primary Study Device: Monarch® eTNS® System

TNS is administered nightly for 4 weeks by the parent during sleep. Stimulation is performed using the Generation 1.0 Monarch**®** eTNS**®** System (NeuroSigma, Inc., Los Angeles), FDA-cleared for unmedicated pediatric ADHD. This device has been used safely and successfully in other prior and ongoing TNS trials at UCLA. The stimulator is worn on the child’s pajamas or T-shirt and is attached with thin wires to disposable, silver-gel, self-adhesive patch electrodes. Parents apply patches across their child’s forehead to provide bilateral stimulation of both V1 trigeminal branches for ∼8 hr nightly. Patches are removed each morning. The active condition uses a 120-Hz repetition frequency, with 250-ms pulse width, and a duty cycle of 30 s on/30 s off. Current settings from 2-4 mA (range 0–10 mA) are established at baseline by titration, which identifies a stimulation level below the participant’s level of discomfort. Power is provided by 9-V Li rechargeable medical-grade batteries (iPower v.3), which are checked and replaced regularly. Families are informed at a scripted presentation that “pulses may come so fast or so slowly that the nerves in the forehead might or might not detect a sensation.” Each night parents turn on the device, press the “up” button until the device reaches the current level set for the patient at titration, pressing “down” to decrease by 0.2-mA steps in case of overshoot. Current to the patch is limited to a safe range (0-10 mA). Some patients may feel some sensation, which generally fades with time.

FDA-clearance for the Monarch eTNS device covers children with ADHD who are not currently taking ADHD medication. PAE does not exclude a child from this labelling but taking ADHD medication does. Our study does admit children who are taking ADHD medication. Our initial assessment found no evidence for additional risks of TNS due to PAE or to concurrent medication. While the initial trials of TNS for ADHD were performed on unmedicated children, the largest trial to date, the ATTENS trial in London [57], explicitly permits concurrent ADHD medication. For other major indications of TNS (depression, epilepsy, migraine) numerous clinical trials have allowed concurrent psychotropic medication without serious adverse events [58]. Therefore, we do not ask parents to take their children off medication. TNS has generally been well tolerated with infrequent and mild side effects across its indications. Nonetheless, we closely monitor for potential side effects and adverse events, including those associated with medication. The UCLA MIRB has classified use of TNS in medicated children and children with PAE in our study as “investigational use of a non-significant risk (NSR) device”. An NSR classification means, “risks no greater than those of everyday life”.

#### TNS Treatment Duration

The treatment duration is set at 4 weeks as this was the optimal time for achieving positive changes in symptoms in the original studies of TNS in pediatric ADHD [36-37]. A follow-up period after 4 weeks is used to measure short-term maintenance of gains. Retention is encouraged by delaying participant compensation until after follow-up.

#### TNS Diary

For each overnight session of TNS, the parent makes an entry into the TNS Diary. Items include start and end times for every day and week of treatment and current level. The purpose is to encourage adherence with the treatment program.

#### TNS Compliance Questionnaire

This questionnaire measures daily compliance with treatment on a daily basis and includes: date, device not worn, stimulation level, time on device, time off device and explanation for any changes in usage not covered by the protocol.

#### Side Effects Rating Scale and Adverse Events Inventory [36]

These instruments assess potential side effects of TNS and adverse events (including serious adverse events) and were used in prior clinical trials of TNS for pediatric ADHD. (Side effects and adverse events were mild and few.) We slightly modified the Side Effects Rating Scale Parent Version to create an additional Side Effects Rating Scale Child Version worded in a way more easily understood by children. Both are completed during Pre-TNS Assessment and at the end of each week of treatment in the presence (telephone, Zoom, or in-person) of a study investigator. The Side Effects Rating Scale Parent is completed by the parent. The Side Effects Rating Scale Child is read to the child by a study investigator. Parents and children are generally advised to report serious emergent side effects or adverse events to the physician investigator as soon as possible for each occurrence.

#### Sleep Quality: Children’s Sleep Habits Questionnaire (CSHQ; [59])

This is a validated, reliable parent-completed test of a child’s sleep quality and sleep behaviors. The CSHQ showed adequate internal consistency for a clinical sample (*p* = 0.78); alpha coefficients for the various subscales of the CSHQ ranged from 0.56 (Parasomnias) to 0.93 (Sleep-Disordered Breathing) for a sleep clinic group. Test-retest reliability was acceptable (0.79). CSHQ individual items, as well as the subscale and total scores consistently differentiated a community group from a sleep-disordered group, demonstrating validity. This measure revealed improvement in sleep with TNS in prior ADHD samples [36-37]. The CSHQ is administered weekly during treatment and at the Pre- and Post-TNS Assessments.

### Pre- and Post-TNS Treatment Assessment Measures

#### ADHD Severity: ADHD-RS

The ADHD Rating Scale (ADHD-RS; [60]) is a parent interview administered by a physician investigator that assesses severity of core symptoms (inattention, hyperactivity) of ADHD. Total Score on the ADHD-RS was the primary outcome in prior clinical trials of TNS for ADHD [36-37]. It is the primary efficacy measure in this trial. It is administered at Pre- and Post-TNS Assessments and at Follow-Up.

#### ADHD Severity: CGi-S/I

The Clinician’s Global Impression Severity (CGI-S) and Clinician’s Global Impression Improvement. (CGI-I; [61]) are secondary efficacy measures in this trial. These extremely widely used, multiply validated 7-point scales assess initial severity, respectively, improvement (or worsening) of core symptoms of a chosen mental disorder (here ADHD) based on a physician’s experience with typical patients. The CGI-S is administered at the Pre-TNS Assessment. The purpose is to serve as one metric of pretreatment severity of ADHD-like symptoms of PAE. The CGI-I is administered at the Post-TNS Assessment and at Follow-Up. The purpose is to serve as a metric of post-treatment change in severity of ADHD symptoms and maintenance of gains at follow-up.

#### ADHD Severity: Conners 4 Parent Short Version (Conners 4 P; [62]

The Conners 4 is a standardized measure that provides a comprehensive assessment of symptoms and impairments associated with ADHD and common co-occurring problems and disorders in children and youth aged 6-18 years. Now fully digital, it provides online scoring for better data visualization, easy inventory management, a digital manual, and printable forms. It also provides a more comprehensive picture of how a rater approaches completing the measure, using updated validity scales, the number of omitted items, and the average number of items completed per minute. The Conners 4 has excellent internal consistency (median omega coefficient = 0.94), strong test-retest reliability (median *r* = 0.89), and convergent validity (median *r* = 0.73). The Conners 4 is administered during the Pre-TNS and Post-TNS Assessments.

#### Vitals

The participant’s vitals (heart rate and blood pressure) are measured at the Pre- and Post-TNS Assessments.

#### Executive Functions: Behavior Rating Inventory of Executive Function (BRIEF II; [63])

The BRIEF II is a standardized validated assessment of executive functions for children and adolescents that is widely used in research. Executive function measures include inhibit, self-monitor, emotional control, initiate behavior, working memory, organization skill, task monitoring, and organization of materials. Summary scores include Behavioral Regulation, Cognitive Regulation, and Global Executive Control. We use the BRIEF II to assess severity of executive dysfunction. Children with ADHD and PAE have been found to have significant problems in executive function on parent ratings of the BRIEF II compared to typically developing controls [64]. The BRIEF II is administered at the Pre- and Post-TNS Assessments.

#### Executive Function: NIH Toolbox Cognition Domain [65]

The NIH Toolbox is a multidimensional set of measures to assess neurological and behavioral function. It is a valid, reliable, brief, and state-of-the-art instrument. We administer the Cognition Domain to our child participants. The Cognition Domain measures basic cognitive and executive functions. The Cognition Domain yields a Cognitive Function Composite, Fluid Cognition Composite (Dimensional Change Card Sort, Flanker Inhibitory Control and Attention, Picture Sequence Memory, List Sorting, and Pattern Comparison measures), and Crystallized Cognition Composite (includes Picture Vocabulary and Reading Recognition measures) Scores. The NIH Toolbox is administered at the Pre- and Post-TNS Assessments.

#### Emotion Regulation: NIH Toolbox Emotion Domain [65]

We administer the Emotion Domain to our child participants and to their parents. The Emotion Domain assesses some aspects of behavioral regulation described in the DSM 5 behavioral phenotype of children with PAE. The Emotion Domain for the child surveys Positive Affect, Emotional Support, Friendship, Loneliness, Perceived Rejection, Perceived Hostility, Self-Efficacy, Sadness, Perceived Stress, Fear, and Anger. The parent version of the Emotion Domain surveys parent impression of the child along the same features. The Toolbox is given at the Pre- and Post-TNS Assessments.

#### Mood: Children’s Depression Inventory 2 (CDI 2; [66])

The Children’s Depression Inventory 2 contains 28 items, each of which consists of three statements. For each item, the child is asked to select the statement that best describes his or her feelings. The CDI 2 assesses self-reported key symptoms of depression, such as feelings of worthlessness and loss of interest in activities, and supports early identification and diagnosis of depressive disorders. Importantly, suicidal ideation as a potential adverse event and a study exclusion criterion is included in the assessment. The CDI 2 is administered at the Pre- and Post-TNS Assessments.

#### Mood: Revised Child Anxiety and Depression Scale Parent and Child Versions (RCADS; [67])

The RCADS assesses both the child report and parent report of youth’s symptoms of anxiety and depression across six subscales: social phobia, panic disorder, major depression, separation anxiety, generalized anxiety, and obsessive compulsive disorder. It also yields a Total Anxiety Scale (sum of the 5 anxiety subscales) and a Total Internalizing Scale (sum of all 6 subscales). One-week test-retest coefficients were good [68] with good concurrent validity compared to the Children’s Depression Inventory and with the Revised Children’s Manifest Anxiety Scale [67]. The RCADS is administered at the Pre- and Post-TNS Assessments.

#### Mood: Affective Reactivity Index Child and Parent (ARI; [69])

The ARI was developed to measure irritability in children from birth to 12 years. The ARI was specifically designed to obtain comparable information from youth and their parents. Using US- and UK- based samples, the parent- and self-report forms of the ARI showed excellent reliability and formed a single factor. In terms of validity, the parent- and self-reported ARI Total Score successfully differentiated cases from controls in clinical and community samples. The parent-rated ARI Total Score also differentiated between youth with severe mood dysregulation and youth with bipolar disorder. In the US sample, Cronbach’s alpha was 0.92 and 0.88, for the parent- and self-report scales, respectively. Regarding construct validity, the ARI showed a gradation with irritability significantly increasing from healthy volunteers through to severe mood dysregulation (SMD). The ARI is administered at the Pre- and Post-TNS Assessments.

#### Magnetic Resonance (MR) Examinations of the Brain

This study explores possible brain bases of TNS therapy and predictors of clinical response. For these purposes, children undergo an ∼75-min examination in a conventional clinical MRI scanner (3 T, 20-channel headcoil). Children are safety screened for MRI contraindications including metal in or on the body or clothing. Children’s heads are firmly and comfortably positioned in the scanner with ample head padding. An MRI-opaque Vitamin E capsule is taped to the right scalp to ensure proper left-right lateralization of images in post-processing. Ear plugs are provided to protect against loud scanner noise. Patient and scanner operator are in audio contact over a loudspeaker and microphone at all times. The patient also has an emergency squeeze bulb to sound an alarm in case of distress. Multiple whole-brain scans are acquired, each in a different MR modality. They include rapid scout structural scans, eyes-open resting-state fMRI (rsfMRI), high-resolution structural MRI, echo-planar spectroscopic imaging (EPSI), and diffusion tensor imaging (DTI). All of these scans are equivalent from a human subjects and safety point-of-view.

#### Parent and Child Satisfaction

Parent and child satisfaction questionnaires are administered at Post-TNS Assessment to measure estimation of the difficulty in administering the treatment, its usefulness for helping the child, overall satisfaction with the outcome, and if parents would recommend the treatment to others.

### Concurrent Medication

Given frequent prescription of psychotropic medication in this population, recruiting exclusively unmedicated children would be impractical. Therefore, medications are allowed in this trial. Their use is included as covariates in statistical analyses. In particular, we control for atomoxetine and guanfacine use in the past two weeks prior to enrollment, as these affect the arousal system thought to be impacted by TNS [57,70]. Patients currently on stimulant medication are asked to withhold medication on assessment days. Otherwise medication continues throughout treatment.

### Safety Monitoring, Adherence, Compliance Monitoring

#### Contraindications to TNS

include metallic implants, e.g., cardiac and neurostimulation systems, and dermatitis or sensitive skin. The Monarch device should not be used in patients with body-worn devices (e.g., insulin pumps or transcutaneous vagal nerve stimulators). There are also hazards with improper use of the device or attaching the patches other than on the forehead. Concurrent use of cellular telephones can interfere with TNS. These TNS risks are mitigated by preselection of patients, by thorough instruction in proper use of the device, and by persistent monitoring for adverse events. For the MRI portion of the study, hazards and safety management are well known and not repeated here.

#### TNS Side Effects and Adverse Events

TNS is generally well-tolerated. Side effects are relatively mild and include: bronchitis, headache, itching, light-headedness, nausea, poor appetite, skin rash, stomach ache, tooth pain, vomiting, trouble sleeping, nightmares, drowsiness, fatigue, tingling sensation, rapid heartbeat, constipation, frequent urination, increased appetite, and teeth clenching. These effects are typically transient. The study device has significant safety data and serious adverse events are not expected, although we do monitor for them. If any unanticipated problems related to the research involving risks to participants or others happen (including serious adverse events) these are reported to the IRB. Adverse events that are not serious but that are notable and could involve risks to participants are summarized in narrative form and submitted to the IRB at the time of continuing review.

During treatment, the parent monitors for side effects and adverse events and completes weekly interim assessments of side effects (Side Effects Rating Scale), adverse-events (Adverse Events Inquiry), and sleep quality (CSHQ). Parents are in regular contact with the study team and are encouraged to report side effects and adverse events promptly. The physician investigator treats or refers such conditions for treatment as needed and decides whether or not discontinuing TNS is called for. These measures mitigate the risk of adverse events which overall are reported as minimal and of low likely impact. The investigators discontinue TNS for any participant who, in the judgement of the study physician, experiences excessive risk (e.g., serious side-effects) or who requires prompt care (e.g., discovery of a serious incidental finding) that is incompatible with continued participation.

#### Strategies to Improve Adherence to Treatment

A key question regarding adherence is whether children will remove the TNS device overnight unbeknownst to their parents. Studies of TNS treatment of pediatric ADHD at UCLA to date are encouraging in that there have not been issues with children removing the device in these studies [36-37]. In principle, however, it could be different for children with ADHD and PAE. In addition to in-person training on the device, parents of each child are given an TNS Instructions and FAQ document at the beginning of treatment. Among several troubleshooting tips, parents are advised what to do if their child removes the device. If they catch it in the course of the night, they are advised to replace the device with fresh electrodes. If they do not become aware until the next morning, they are advised to note the incident in the TNS Diary and to contact the study team. When reapplying the device the next evening, parents are advised that putting a headband over the electrodes may help, especially if the child is a restless sleeper. Additionally, the parents are advised to ensure that no lotions or creams, that may facilitate slippage, are applied to the skin under or around the electrode.

#### Compliance Monitoring

TNS “compliance” is defined as the child wearing the device for 21 of 28 nights of treatment. The parent maintains a TNS Diary to ensure compliance, reinforced by daily telephone contact with investigative staff. Non-compliance is failure to wear the device for 7 hours or more on 8 or more nights during the treatment regimen. In such cases, we contemplate investigator-withdrawal of the child from the study. In the TNS Instructions and FAQ document and on the ICF, parents are informed that their child may be withdrawn from the study if he or she repeatedly removes the device overnight or otherwise fails to comply with study procedures. For participants who are withdrawn, all within-compliance data collected up to the time of withdrawal are retained and no further data are collected after withdrawal.

### Statistical Analysis

#### Primary Objective

The main objective of this study is to evaluate whether TNS treatment of ADHD in PAE is feasible under the conditions described. “Feasible” for this purpose means “safe, well tolerated, and promising efficacy”. The corresponding criteria are **Safety:** 1) No meaningful (i.e., serious morbidity or mortality) TNS-related adverse events; 2) Side effects requiring extensive remediation or interfering with study procedures in #x2264;3 patients (10%); **Tolerability:** 3) #x2264;3 patients (10%) withdrawing from the study based on side effects or poor tolerance of TNS; 4) TNS compliance ≥21/28 sessions (75%) in ≥24/30 patients (80%) (based on TNS diaries); **Promise of Efficacy:** 5) ≥25% reduction in ADHD-RS and/or CGI-I of 2 or 1 in 210 patients (33.3%). If these criteria are met, genuine efficacy can then be tested more rigorously in a future double-blind, sham-controlled RCT.

#### Secondary Objectives: Behavioral

Ancillary objectives include determining whether TNS treatment improves executive function, mood, and/or sleep. These will be tested by comparing Post-TNS to Pre-TNS scores on endpoints from the BRIEF II, Conners 4, NIH Toolbox, CDI 2, RCAD, ARI and CSHQ and looking for pre and post treatment significant differences. In addition, we will determine whether or not the participant moves from scores in the clinical range on standardized measures to the nonclinical range following treatment.

#### Secondary Objectives: MR Neuroimaging

A final set of objectives is pursued by way of multimodal MRI measurements, including structural MRI, MRS, DTI, and rsfMRI. Briefly, we will test hypotheses regarding MR endpoints in anterior cingulate cortex, middle frontal cortex and inferior frontal cortex. These are brain regions which both show abnormalities in PAE and appear to be affected by TNS. The endpoints include local gyrification index (LGI; cortical curvature) and cortical myelin content measured by MRI, glutamate (Glu) concentration measured by MRS, mean diffusivity measured by DTI, and functional connectivity measured by rsfMRI. We hypothesize that group-mean Glu will increase and functional connectivity will decrease in these regions following TNS. We further hypothesize that low pre-TNS Glu, LGI and myelin and high MD and functional connectivity predict favorable post-TNS clinical response.

#### Data Management

Data are entered into an electronic database managed by the UCLA Semel Institute Biostatistics Core. Procedures include scanning for duplicate entries and range checks for variable values. Personal health information on participants is not shared outside the study. All members of the study team are certified in participant confidentiality. Paper records are kept in a locked filing cabinet; electronic records are stored on password-protected media.

#### General Statistical Procedures

Mean differences and predictive associations are tested using the Linear Mixed Model in R with covariates and with last observation carried forward (LOCF) for early withdrawal patients. Covariates include demographics (sex, age, ethnicity), IQ Composite on the KBIT-2, maternal education (an established proxy for socioeconomic status), other teratogen exposures (smoking, marijuana, illegal and prescription medications, caffeine), and clinical variables (DSM-5 comorbidities, child current medications). Possible covariates that are associated with outcomes at *p* < 0.10 will be included in final models. No correction for multiple comparisons is made for comparisons with *a priori* hypotheses. For other tests, FDR is applied. Efficacy is computed twice. Efficacy is computed once including all patients, both those who complete the study and those who were withdrawn using LOCF; efficacy is computed again including only completers. Both computations are useful and relevant. The former mitigates against bias in rejecting non-responders. The latter tells what efficacy is if compliance is achieved, a reasonable goal in most cases.

## Discussion

This study protocol is for an open-label pilot trial seeking to establish the feasibility (safety, tolerability, promise of efficacy) of 4 weeks of nightly TNS in improving ADHD symptoms in children with prenatal alcohol exposure (PAE) and whether effects persist at 4-week follow-up. Secondary aims are to test whether TNS improves executive function, mood, and sleep quality. We further explore possible predictors of response and brain mechanisms of action in PAE using morphometric MRI, DTI, MRS, and rsfMRI.

Given prior trials in children with ADHD not evaluated for PAE [36-37], we expect to replicate clinical efficacy on ADHD symptoms with medium effect size in children with PAE. This would be highly relevant as first-line treatment for ADHD (psychostimulants) is often ineffective in PAE and/or induces untoward effects. If TNS is safe and efficacious, then a novel non-pharmacological treatment could become available for PAE. Other non-pharmacological treatments for ADHD symptoms, such as behavioral therapies, cognitive training [71], EEG neurofeedback [72], transcranial direct current stimulation (tDCS), fMRI neurofeedback, or dietary interventions, have only small to moderate efficacy. TNS is the non-pharmacological treatment with the largest effect size so far in improving ADHD and is the only non-pharmacological medical device licensed as treatment for ADHD by the FDA.

A prior trial of TNS showed improvements in executive functions in ADHD [36]. We therefore expect the present study to show improvements in executive functions in children with PAE by using a battery of ADHD-relevant instruments.

Like most treatments, TNS is likely to improve ADHD symptoms in a particular subgroup of youth with ADHD. It will be crucial to understand which clinical, cognitive, or neuroimaging factors predict treatment response. For this purpose, we will test for treatment response prediction using pre-TNS values of clinical, cognitive and MR measures and have cast hypotheses accordingly.

Currently knowledge of how TNS works in the brain in ADHD or any other condition is incomplete [58]. Rubia et al. [57] detail a plausible mechanism of action for improvement of attention in ADHD. Briefly, TNS electric stimulation is conveyed from the face over the trigeminal nerves to multiple subcortical sites, including nucleus solitarius, locus coeruleus, and the reticular activation system. Through adrenergic projections and modulation of dopamine and Glu levels in cerebral cortical target regions [73-74], stimulation of the locus coeruleus and the reticular activation system is especially effective in increasing arousal, inhibiting distraction, facilitating task-relevant responses, improving sustained attention and vigilance and thereby enhancing cognitive performance. The multimodal MR investigations in this protocol will examine relationships between regional brain structure, metabolism, and functional connectivity and TNS clinical responses. This may aid in deciphering brain mechanisms of TNS in children with ADHD and PAE. Thus, this study may both introduce a novel non-pharmacological treatment for ADHD in PAE with minimal side effects and provide a rational basis for that therapy.

## Supporting information

SPIRIT Chiecklist

CONSORT Diagram

## Supplemental materials

**SM1 SPIRIT checklist**

**SM2 CONSORT Diagram**

## Acknowledgments

This research is supported by a grant R61AA031029 (Joseph O’Neill, PI) from the National Institute on Alcohol Abuse and Alcoholism Number, clinical trial #NCT06847165. The content is solely the responsibility of the authors and does not necessarily represent the official views of the National Institute on Alcohol Abuse and Alcoholism or the National Institutes of Health. The funders had no role in study design, data collection and analysis, decision to publish, or preparation of the manuscript.

## Data availability statement

Deidentified research data will be made publicly available in the NIAAA Data Archive after the study is complete. Results will be disseminated by publication.

## Ethics approval

All procedures to be performed involving human participants are in accordance with the ethical standards of the UCLA MIRB and with the 1964 Helsinki declaration and its later amendments or comparable ethical standards. This study was approved by the UCLA Medical Institutional Review Board (MIRB).

## Consent to participate

The UCLA Medical Institutional Review Board approved all procedures. All procedures are in accordance with the ethical standards of the institutional and/or national research committee and with the 1964 Helsinki declaration and its later amendments or comparable ethical standards. Parents and children give their informed consent/assent prior to study inclusion.

## Author contributions

**Conceptualization:** Joseph O’Neill and Mary J. O’Connor

**Funding acquisition:** Joseph O’Neill

**Methodology:** Joseph O’Neill, Mary J. O’Connor, Jeffrey R. Alger and Shantanu H. Joshi

**Project administration:** Joseph O’Neill

**Writing original draft:** Joseph O’Neill

**Writing –review & editing:** Joseph O’Neill, Mary J. O’Connor, Jeffrey R. Alger and Shantanu H. Joshi, Benjamin N. Schneider

All authors read and approved the final manuscript.

