## Supplementary material for "Trigeminal Nerve Stimulation (TNS) for Children with Attention Deficit/Hyperactivity Disorder and Fetal Alcohol Spectrum Disorder: Feasibility Study Protocol": CONSORT Diagram

Template CONSORT diagram

**Included in ITT analysis (n=)**

Interested/Pre-screened (n=)

Excluded (n=)

Reasons (n=)

Written consent (n=)

Excluded (n=)

Reasons (n=)

Completed Eligibility Assessment (n=)

Excluded (n=)

Reasons (n=)

Completed Pre-TNS Assessment (n=)

Allocated to TNS (n=)

Excluded (n=)

Reasons (n=)

Discontinued treatment (n=)

Reasons (n=)

Completed TNS treatment and Post-TNS Assessment (Primary Endpoint)

(n=)

*n= missing data*

*Withdrawn from data collection (n=)*

*Reasons (n=)*

Completed 4-week Follow-Up (n=)

*n= missing data*

*Withdrawn from data collection (n=)*

*Reasons (n=)*
